# How do Hemodialysis Center Prevent the Secondary COVID-19 Transmission in Poverty-Stricken Rural Region of China: Experiences and Strategies

**DOI:** 10.1101/2021.03.09.21252639

**Authors:** Yue Gu, Yuming Wang, Jing Zhou, Yaorui Deng, Fengmin Shao

**Affiliations:** Department of Nephrology, Henan Provincial People’s Hospital and the People’s Hospital of Zhengzhou University, Zhengzhou, China; Clinical Research Center, Henan Provincial People’s Hospital and the People’s Hospital of Zhengzhou University, Zhengzhou, China; Department of the Health Management, Henan Provincial People’s Hospital and the People’s Hospital of Henan University, Zhengzhou, China; Hemodialysis Center, Huangchuan County People’s Hospital, Xinyang, China

## Abstract

The COVID-19 pandemic has caused an evolving public health crisis and challenged the medical system globally, especially in the rural-stricken regions. There is concern about the spread of coronavirus in regions with lower education level, weaker health systems and underdeveloped economy. The risk of viral transmission in HD center is elevated because of the densely-populated and high mobility in an enclosed environment.This paper demonstrated the main experiences and strategies of preventing secondary COVID-19 transmission in a HD center from a poverty-stricken rural region in China. Data of subjects including 17 medical workers and 249 patients were collected from the HD center in Huangchuan County People’s Hospital from February to April 2020. It is the first paper to provide the experiences and strategies about preventing COVID-19 secondary transmission in HD center for poverty-stricken rural region.

## Introduction

The novel coronavirus disease (COVID-19) pandemic has caused an evolving public health crisis and challenged the medical system globally, especially in poverty-stricken region (1). The susceptibility of COVID-19 infection in hemodialysis (HD) patients is increased because that they are of older age and have other health issues including decreased immune function, diabetes, hypertension lung diseases and so on. As the COVID-19 is insidious and highly infectious, it is prone to cause cluster transmission. There is increased risk of viral transmission in HD center as a densely-populated, high mobility and enclosed environment. There was a COVID-19 outbreak occurred in the HD center of Renmin Hospital, Wuhan University (2).

This study demonstrated the experiences and strategies of preventing secondary COVID-19 transmission in a HD center from a poverty-stricken rural region in China.

## Methods

Data of subjects including 17 medical workers and 249 patients were collected from the HD center in Huangchuan County People’s Hospital from February to April 2020. The study was approved by the local institutional review board, and informed consents were obtained from all participants.

All subjects underwent chest computed tomographic (CT) scans and hematological examination for screening. Real-time reverse transcriptase polymerase chain reaction (RT-PCR) for SAR-CoV-2 detection were performed using nasopharyngeal swabs for suspected cases.

## Results

The HD center had actively screened all patients for potential exposure to confirmed/suspected COVID-19 case daily according to the information from local Center for Disease Control and Prevention. On day 1, patient 1 was found that lives in the same village with the new confirmed case on that day and closely contacted with the people who was exposed to the confirmed case. Patient 1 had mild cough, no elevated temperature, no sore throat or gastrointestinal symptom, and was informed for chest CT and hematological examination. On day 2, patient 1 was diagnosed as a suspected case according to the COVID-19 Diagnosis and Treatment Guideline (modified 5^th^ version) (3) and admitted to the isolation ward. This patient had RT-PCR test result positive for COVID-19 within 1 day. Patient 1 had negative RT-PCR test results and relieved symptoms after treatment, and then was observed and received HD in isolation ward for another 14 days.

Patient 2 through 4 were suspected cases and admitted to the isolation ward for HD for 14 days. The RT-PCR tests were negative. On day 2, patients in adjacent beds of patient 1 within the past 14 days were actively screened. On day 3, patients 2 and 3 were diagnosed as suspected cases. From day 4 to day 7, all of the HD patients, medical workers, accompanying family members and cleaners were then screened, and patient 4 was diagnosed as suspected case.

On day 2, all of the staff in the HD center were screened and isolated from their families in a hotel for 2 weeks. Their daily work continues normally.

The characteristics of the study cohort were summarized in Table 1.The main preventive experience and strategies of HD center during the COVID-19 epidemic are shown in table 2. It has been reported that there was no secondary COVID-19 transmission in HD center.

**Table 1.** Demographic Characteristics of Patients and Medical Workers.

| Characteristics | Patients<br>(n=249) | Medical workers<br>(n=17) |
| --- | --- | --- |
| HD duration, median (IQR), months | 39(20-72) | NA |
| Sex |  |  |
| Female, No. (%) | 102(40.9%) | 14(82.4%) |
| Male, No. (%) | 147(59.1%) | 3(17.6%) |
| Degree of education |  |  |
| Completed primary education, No. (%) | 161(64.7%) | 0 |
| Completed secondary education, No. (%) | 83(33.3%) | 0 |
| Completed diploma, No. (%) | 5(2.0%) | 16(94.1%) |
| Completed bachelor degree or above, No. (%) | 0 | 1(5.9%) |
| Abbreviations: NA, not applicable. |  |  |

**Table 2.** Main Experiences and Strategies for Preventing Secondary COVID-19 Transmission in HD Center.

| Main experiences/strategies | Practice content |
| --- | --- |
| Training | Objects: medical workers, HD patients, accompanying family members and cleaners.<br>Platform: Wechat, Welink and other online teaching channel.<br>Content: 1. COVID-19 knowledge, hand hygiene, mask wearing, respiratory etiquette, social distance, avoid public transportation, temperature measurement, PPE, mental health, environmental decontamination;<br>2. Patients and accompanying family members wear masks during HD;<br>3. Accompanying family members are restricted. |
| Active screening | 1. Work with the local CDC for daily screening of HD patients who is potential exposed to confirmed/suspected case, and then inform them to get relevant test as soon as possible;<br>2. Before entering the HD section, the patients' temperature are measured, ask if there is potential exposure to COVID-19 case and any respiratory or gastrointestinal symptoms. |
| Management of medical workers after finding confirmed/suspected case | 1. Medical workers and cleaners are isolated from their families in the hotel for 14 days;<br>2. PPE in medical workers during the treatment of confirmed/suspected COVID-19 HD patients: goggle, medical isolation suit, isolation gown, N95 respirator and surgical mask, surgical glove, surgical cap, rubber shoes, shoe covers;<br>3. PPE in medical workers during the treatment for non-confirmed/non-suspected COVID-19 HD patients: face screen, isolation gown, surgical mask, latex glove, surgical cap, shoe covers. |
| Management of confirmed/suspected case | 1. The confirmed COVID-19 HD patient is treated in isolation ward. After RT-PCR test is negative and the symptoms is relieved, patient then is observed and received HD in isolation ward for another 14 days;<br>2. The suspected case is observed and received HD in isolation ward for 14 days. |
| Management of other patients after finding confirmed/suspected case | 1. For the HD patients who was in the adjacent bed of confirmed/suspected case within the past 14 days, the relevant test are carried out as soon as possible. If it's necessary, all patients in HD center are screened;<br>2. Self isolation at home during non-HD time;<br>3. Reduce the hospital detention time, and the time of non-HD in the hospital is less than 30 minutes. |
| Abbreviations: HD, hemodialysis; PPE, personal protective equipment; CDC, Center for Disease Control and Prevention; RT-PCR, real-time reverse transcriptase polymerase chain reaction. |  |

## Discussion

As a poverty-stricken rural region, the per capita disposable income in the reported area was 1,962 USD (4). The described HD center is the only HD center in this area. Even though the education level in medical workers and HD patients is low, the active screening in HD center from this hospital prevented the secondary COVID-19 transmission. Similarly, during the 2015 MERS-CoV outbreak in Korea, although 116 patients were exposed to MERS-CoV in HD units, but strict patient surveillance and appropriate isolation measures prevented secondary transmissions (5). In poverty-stricken rural region, the lack of protective awareness in asymptomatic carriers or patients with mild symptoms due to the low education level might lead to the further spread of coronavirus which can cause overwhelmed local medical system. Therefore, the implementation of active screening and identification of COVID-19 confirmed/suspected case in HD center is important in poverty-stricken rural regions.

This study was limited as a single-centered study with small sample size. Future multi-centered studies are needed for establish strategies for preventing secondary transmission in HD center from poverty-stricken rural region.

## Data Availability

All data and information described in the paper is open to public.

